# Treatment of Moderate to Severe Respiratory COVID-19—A Cost-Utility Analysis

**DOI:** 10.1101/2020.09.21.20199182

**Authors:** Stephen E. Congly, Rhea A. Varughese, Crystal E. Brown, Fiona M. Clement, Lynora Saxinger

**Affiliations:** Division of Gastroenterology and Hepatology, Department of Medicine, Cumming School of Medicine, University of Calgary, Calgary AB, Canada; O’Brien Institute of Public Health, University of Calgary, Calgary AB, Canada; Division of Pulmonary Medicine, Department of Medicine, University of Alberta, Edmonton AB, Canada; Division of Pulmonary, Critical Care and Sleep Medicine, University of Washington, Seattle WA, USA; Department of Community Health Sciences, University of Calgary, Calgary AB, Canada; Division of Infectious Diseases, Department of Medicine, University of Alberta, Edmonton AB, Canada

## Abstract

**Background:** Due to COVID-19’s significant morbidity and mortality, identifying the most cost-effective pharmacologic treatment strategy is critical. As such, we determined the most cost-effective strategy for moderate to severe COVID-19 respiratory infections using the United States health care system as a representative model.

**Methods:** A decision analytic model modelled a base case scenario of a 60-year-old patient admitted to hospital with COVID-19. Patients requiring oxygen were considered moderate severity, and patients with severe COVID-19 required intubation with intensive care. Strategies modelled included giving remdesivir to all patients, remdesivir in severe infections, remdesivir in moderate infections, dexamethasone to all patients, dexamethasone in severe infections, remdesivir in moderate/dexamethasone in severe infections, and best supportive care. Data for the model came from the published literature. The time horizon was 1 year; no discounting was performed due to the short duration. The perspective was of the payer in the United States health care system.

**Results:** Supportive care for moderate/severe COVID-19 cost $11,112.98/0.8256 QALY. Remdesivir in moderate/dexamethasone in severe infections was the most cost-effective with an incremental cost-effectiveness ratio of $19,764.56/QALY gained compared to supportive care. Probabilistic sensitivity analyses showed remdesivir for moderate/dexamethasone for severe COVID-19 infection was most cost-effective in 88.6% of scenarios and dexamethasone in moderate-severe infections in 11.4% of scenarios. With lower willingness to pay thresholds ($250-$37,500), dexamethasone for severe infections was favoured.

**Conclusions:** Remdesivir for moderate/dexamethasone for severe COVID-19 infections was the0020most cost-effective strategy. Further data is required for remdesivir to better assess its cost effectiveness in treatment of COVID-19.

## Introduction

Severe acute respiratory syndrome coronavirus 2 (SARS-CoV-2) was first reported to cause respiratory illness in China in December 2019, with the disease designated as COronaVIrus Disease 2019 (COVID-19)^1^. SARS-CoV-2 has caused significant morbidity and mortality globally; as of September 21, 2020, there have been 31,200,220 documented cases worldwide and 963,000 confirmed deaths with the highest national burden thus far in the United States^2^. Given this impact, there has been great interest in finding potential treatments for COVID-19^3^ although early enthusiasm and adoption of possible candidate drugs, such as hydroxychloroquine, has been tempered upon rigorous study^4^.

Currently, two drugs have shown benefit in randomized controlled trials. First, remdesivir, a RNA polymerase inhibitor with broad antiviral activity and *in vitro* effect against SARS-CoV-2^3^, was shown to have benefit in a randomized controlled trial in the United States sponsored by the National Institute of Health^5^ with a list price of $2340 US dollars per treatment course^6^. More recently, dexamethasone was shown to improve COVID-19 outcomes in a randomized controlled trial in the United Kingdom^7^ with a list price of approximately $20 per treatment course^8^. Incomplete 28-day mortality data suggests that the benefit of antiviral therapy with remdesivir is highest in patients needing supplemental oxygen with a 8.5% reduction in mortality and a reduced length of stay^5^, while the anti-inflammatory effect of dexamethasone appears somewhat beneficial in patients on oxygen, reducing mortality by 3.5%, and most beneficial in those requiring mechanical ventilation with a reduction of mortality by 11.7%^7^. Given the economic implications of COVID-19 and these potential treatments, we developed a cost-effectiveness analysis of these drugs in the United States context with additional global considerations assessed by willingness-to-pay thresholds.

## Methods

### Model Design

A decision tree was developed with TreeAge Pro 2020 (TreeAge Software, Williamstown MA) to create a cost-effectiveness analysis for patients with moderate-severe COVID-19 respiratory infections. The base-case scenario was a 60-year-old patient admitted to hospital with a respiratory COVID-19 infection. Cases could either be admitted to the ward on oxygen (moderate COVID-19 infection) or the intensive care unit (ICU) (severe COVID-19 infection). The strategies compared were chosen based on current published data: giving dexamethasone to all patients, dexamethasone to only severe COVID-19 infections, remdesivir to all patients, remdesivir to moderate COVID-19 infections only, remdesivir to only severe COVID-19 infections, remdesivir to moderate and dexamethasone to severe COVID-19 infections, and best supportive care to all patients. Patients who did not receive either therapy were assumed to receive best supportive care. For patients receiving dexamethasone, they were assumed to receive 6 mg per day for 10 days^7^. For patients receiving remdesivir for severe COVID-19 infections, they were assumed to receive 200 mg on day 1 and then 100 mg daily for 9 additional days. For moderate COVID-19 infections, patients were assumed to receive a 5 day course of remdesivir; 200 mg on day 1 and then 100 mg daily for 4 additional days given data suggesting similar efficacy of 5 and 10 day courses in this population^9^. In this simple model, a patient would either recover from their infection and survive or die with no transition between ward and ICU. To account for clinical situations in which patients may be admitted to the ICU on high flow oxygen, a scenario analysis was performed where all patients were assumed to be admitted to the ICU with the expected increase in costs. It was assumed that beyond 28 days, there would be no ongoing conditions expected to significantly impact assessed quality adjusted life years for the rest of the time-horizon.

### Data Sources

Data for the model come from the published randomized controlled trial literature^5,7,9^. Hospital costs were based on the 2020 Medicare national payment rate^10^ for the appropriate diagnosis related group (DRG) code with moderate COVID-19 respiratory infections being classified as DRG 178 and severe COVID-19 infections classified as DRG 207. Prices are based on the 2020 Medicare Part B database^8^ for dexamethasone and the published price for remdesivir^6^. Utilities were based on previous experiences with H1N1 and influenza^11,12^; patients were assumed to have these utilities for 28 days and then return to the US average utility of 0.947^13^ for the remainder of the year if they survived. Costs are presented in 2020 US dollars. Model inputs are listed in Table 1.

**Table 1:** Model Inputs

| Variable | Variable | Range Tested | Distribution for Multivariate Analysis | Reference |
| --- | --- | --- | --- | --- |
| Probability of having severe COVID-19 | 0.15 | 0.05-0.25 | Beta | <sup>5,7</sup> |
| Probability of death with severe COVID-19 | 0.25 | 0.15-0.45 | Beta | <sup>5,7</sup> |
| Probability of death with moderate COVID-19 | 0.07 | 0.04-0.20 | Beta | <sup>5,7</sup> |
| Risk reduction of death with dexamethasone for severe COVID-19 | 0.65 | 0.5-0.82 | Log-normal | <sup>7</sup> |
| Risk reduction of death with dexamethasone for moderate COVID-19 | 0.8 | 0.70-0.92 | Log-normal | <sup>7</sup> |
| Risk reduction of death with remdesivir for severe COVID-19 | 1.08 | 0.91-1.28 | Log-normal | <sup>5</sup> |
| Risk reduction of death with remdesivir for moderate COVID-19 | 0.29 | 0.20-0.42 | Log-normal | <sup>5</sup> |
| Cost dexamethasone for 10-day course at 6 mg/day | 19.20 | 15-30 | Gamma | <sup>8</sup> |
| Cost remdesivir per day | 234 | 100-350 | Gamma | <sup>6</sup> |
| Cost severe COVID-19 admission [USD] [DRG 207] | 33,247.15 | 14400-50000 | Gamma | <sup>10</sup> |
| Cost moderate COVID-19 admission [USD] [DRG 178] | 7206.95 | 5020.46-10962.59 | Gamma | <sup>10</sup> |
| Utility severe COVID-19 | 0.23 | 0.18-0.28 | Beta | <sup>12</sup> |
| Utility moderate COVID-19 | 0.5616 | 0.3846-0.6925 | Beta | <sup>11</sup> |

### Methodology

The perspective of the health care payer was utilized with a willingness to pay threshold of $100,000/quality adjusted life year (QALY). A 1-year time horizon was used in this model; discounting was not done due to the short time frame. Univariate analysis and a multivariate probabilistic sensitivity analysis using 100,000 simulations were performed to assess the model. The model was internally validated and was determined to have good face validity. We followed the CHEERS checklist when writing our manuscript^14^ and our model was in keeping with best practice guidelines^15^. Ethics approval was not required as the data used in this study came from publicly available data.

## Results

In the base-case model (Figure 1, Table 2), supportive care for moderate-severe COVID-19 had a cost of $11,112.98 for 0.8256 quality adjusted life years (QALY). Using remdesivir for moderate and dexamethasone for severe infections had an incremental cost effectiveness ratio (ICER) of $19,764.56/QALY versus supportive care and was the most cost effective of the strategies evaluated. Using remdesivir for all patients or remdesivir for severe infections only were both less effective and more costly than dexamethasone-based strategies and were considered dominated. Using remdesivir for moderate COVID-19 infection only was considered to be extendedly dominated, meaning that the incremental cost-effectiveness ratio was higher than a more effective strategy.

**Table 2:** Base Case Analysis Referencing Supportive Care as Baseline

| **Strategy** | **Cost [US\$]** | **Incremental Cost [US\$]** | **Efficacy [QALY]** | **Incremental Efficacy [QALY]** | **Incremental Cost Effectiveness Ratio [US\$/QALY]** |
| --- | --- | --- | --- | --- | --- |
| Supportive care | 11,112.98 | - | 0.8256 | - | - |
| Dexamethasone severe | 11,115.86 | 2.88 | 0.8373 | 0.0117 | 246.00 |
| Dexamethasone all | 11,132.18 | 19.20 | 0.8482 | 0.0226 | 848.64 |
| Remdesivir severe | 11,463.98 | 351.00 | 0.8229 | -0.0027 | DOMINATED |
| Remdesivir moderate | 12,107.48 | 994.50 | 0.8643 | 0.0388 | EXTENDEDLY DOMINATED |
| Remdesivir moderate, dexamethasone severe | 12,110.36 | 997.38 | 0.8761 | 0.0505 | 19,764.56 |
| Remdesivir all | 12,458.48 | 1345.50 | 0.8617 | 0.0361 | DOMINATED |

**Figure 1:**
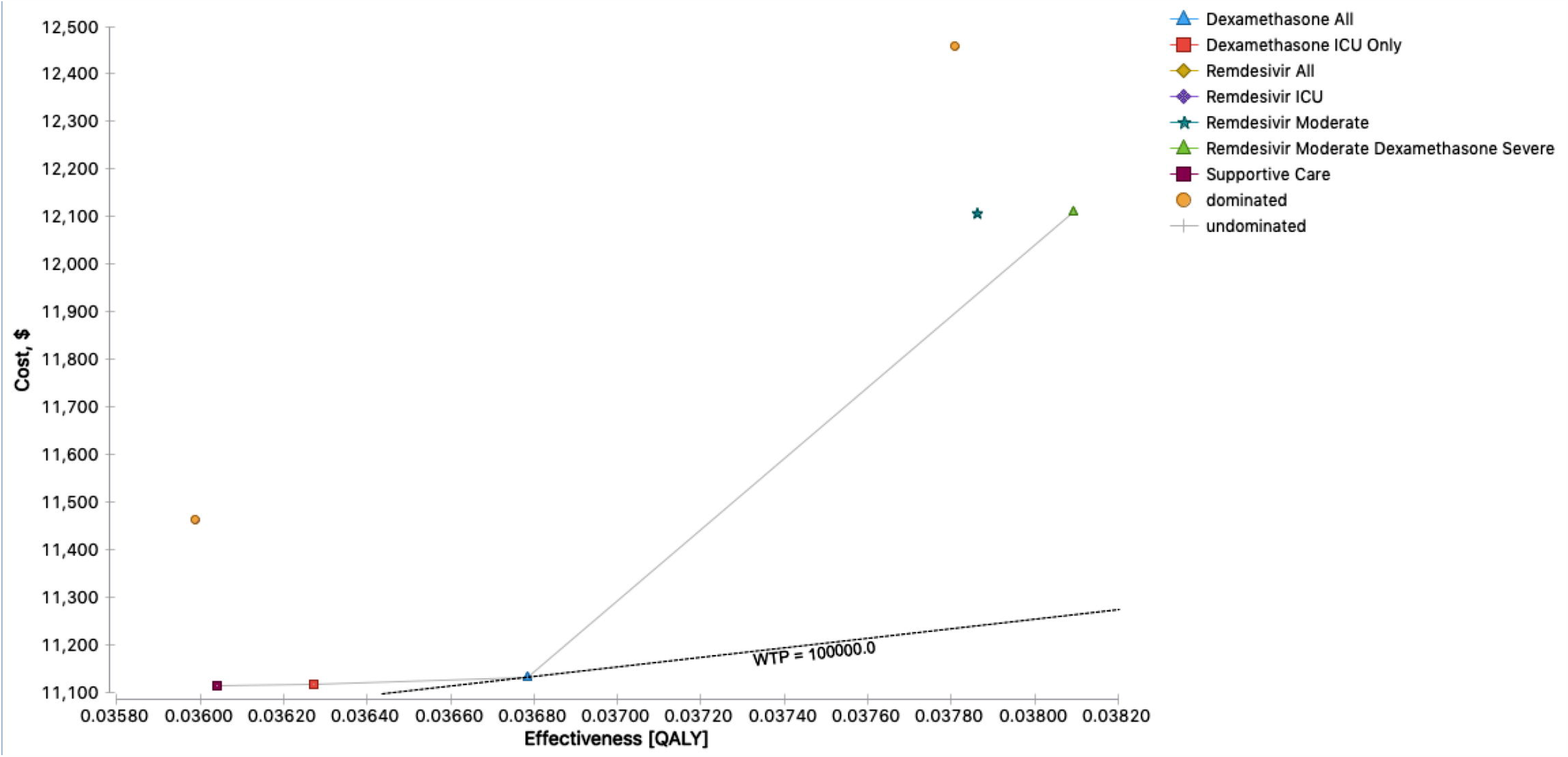
Cost-Effectiveness of Treatments for Moderate-Severe Respiratory COVID-19 Infections. Comparison of all strategies modelled for moderate-severe respiratory COVID-19 infections. The remdesivir for all and remdesivir for severe infections strategies are dominated (less effective and more costly). Remdesivir for moderate infections is extendedly dominated meaning that the incremental cost-effectiveness ratio was higher than a more effective strategy. WTP = Willingness to pay threshold. QALY = Quality adjusted life year.

In the scenario analysis where all patients were assumed to be admitted to the ICU, supportive care now cost $33,247.15/0.8256 QALY. The incremental cost-effectiveness ratios and rankings remained unchanged from the base case except that each strategy cost an additional $22,134.17 accounting for the additional costs of ICU admission as compared to admission to the ward.

In univariate sensitivity analysis, the preferred strategy of remdesivir for moderate and dexamethasone for severe COVID-19 infections remained unchanged with all variables. The ICER for the remdesivir for moderate strategy exceeded $100,000 if remdesivir cost >$4.08 per tablet, the probability of death of moderate COVID-19 was >0.055, the probability of death of severe COVID-19 was >0.386 and the risk reduction of remdesivir for moderate infections >0.41. Remdesivir for moderate and severe COVID-19 infections was dominated in all cases (more expensive and less effective) unless the cost per tablet was less than $0.30.

Probabilistic sensitivity analysis of all strategies based on a willingness to pay threshold of $100,000 showed that the use of remdesivir in moderate and dexamethasone in severe COVID-19 infections would be favoured in 88.58% of simulations while dexamethasone in moderate-severe COVID-19 infections would be the most cost-effective strategy in 11.38% of simulations. All other strategies were extremely unlikely to be cost-effective with probabilities between 0.001-0.002%.

The cost effectiveness acceptability curve (Figure 2) shows that as the willingness to pay threshold increases the remdesivir for moderate and dexamethasone for severe infection strategy continued to be more likely to be favoured. Conversely, with willingness to pay thresholds lower than the typical US standard of $100,000, the use of dexamethasone for all hospitalized infections would be favoured with a willingness to pay between approximately $250-$37,500; supportive care would be favoured with a willingness to pay threshold of less than $250.

**Figure 2:**
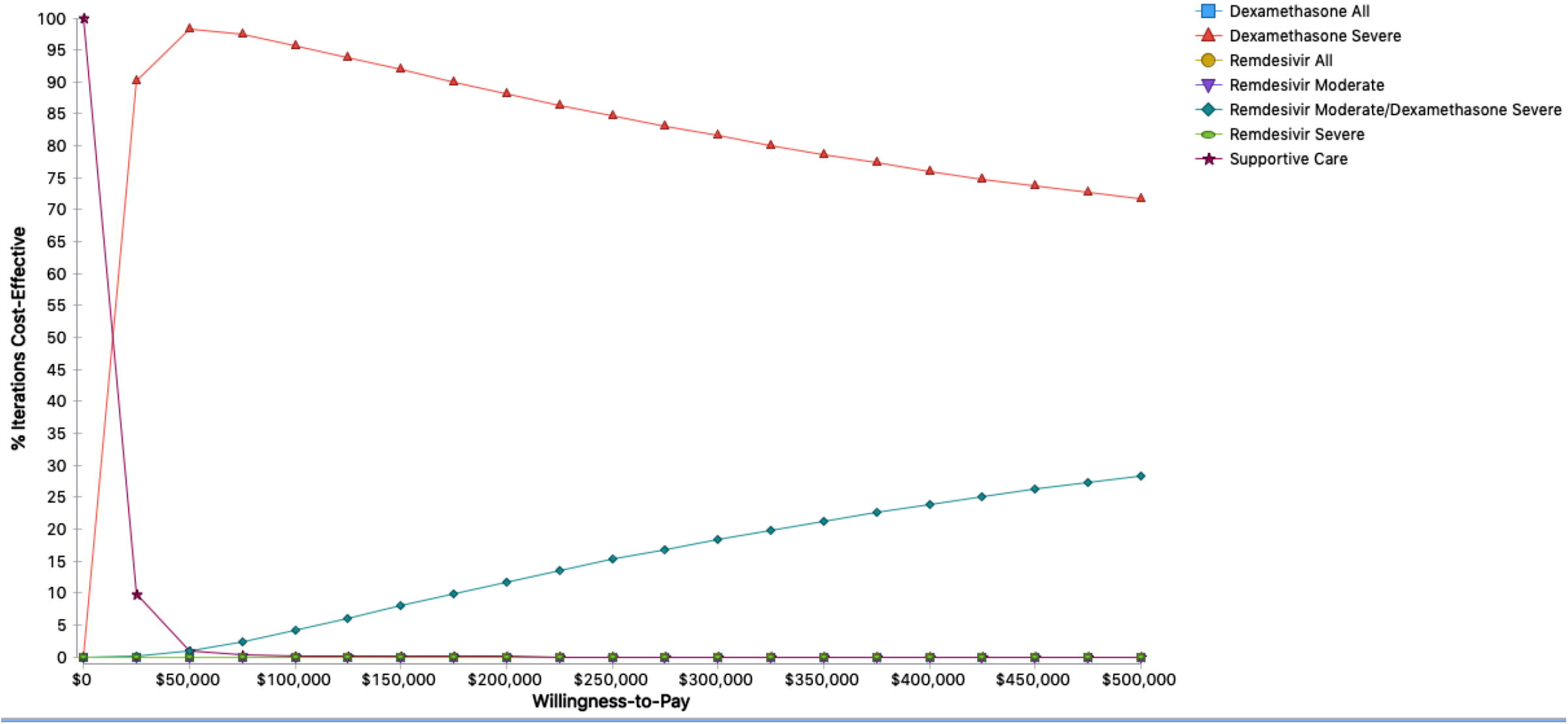
Cost Effectiveness Acceptability Curve with Probabilistic Sensitivity Analysis. Analysis of likelihood of a strategy being preferred based on the willingness to pay threshold in 2020 US dollars. Dexamethasone for severe infections, remdesivir for moderate infection and remdesivir for all overlap at bottom.

## Discussion

In our base-case analysis, we found that using remdesivir for moderate and dexamethasone for severe infections was the most cost-effective strategy to treat moderate-severe COVID-19 infections with a cost of $13,823/QALY. No strategies using remdesivir monotherapy were favoured and in most cases were less effective and more costly than other strategies in the base-case. Multivariate analysis showed that remdesivir for moderate and dexamethasone for severe infections remained the preferred choice when willingness to pay thresholds are over $37,500 USD, which is a relevant threshold in countries such as Canada, Ireland, Australia, Spain and the United Kingdom^16^.

Given the significant morbidity and mortality of moderate-severe COVID-19 infection, development of a vaccine will be critical. In the interim, any treatment that may improve outcomes is valuable but must be balanced with treatment affordability and health care sustainability given the opportunity cost associated with use of these drugs. This model compares the two agents with current randomized controlled trial data showing potential mortality benefit in moderate-severe COVID-19 infections, with remdesivir showing a statistical trend to survival in the preliminary report with a larger effect in patients with moderate infections and dexamethasone demonstrating a statistically significant benefit in both moderate and severe infections. Combining remdesivir for moderate infections and dexamethasone for the most severe infections was found to be the most effective and economical strategy with willingness to pay thresholds over $37,500 USD.

The model design uses a fixed cost for admission with the DRG code and does not account for potential shorter stays in hospital. Given our use of the payer’s perspective, the cost of the hospital stay will be the same rate based on the DRG code regardless of the length of stay. In our base case scenario, we assumed that moderate COVID-19 infections would be admitted to the ward and severe infections admitted to the ICU. This practice is not consistent throughout the United States; some centers will admit patients on high flow oxygen to ICU. In our analysis of this scenario, we found that the league table and ICER values between strategies remain unchanged, but all strategies would cost an additional $22,134.17.

The economic and health impact of COVID-19 has been substantial globally^17^; treatments that can reduce its burden are eagerly sought. Use of either dexamethasone or remdesivir for COVID-19 needs to be considered in the context of the local burden of COVID-19 disease as well as healthcare budgets and priorities. Decisions of treatment and resource utilization need to be made rationally and with consideration of the values and priorities of the population. Economic analysis with cost/QALY can help optimize resource utilization and to try to promote equity for accessing treatment of diseases^18^. Although our representative analysis utilized a United States perspective, we feel that these results can be extrapolated to other jurisdictions world-wide, as the major cost for each strategy was hospitalization; with lower costs of hospitalization, the incremental cost-effectiveness ratios between choices will remain similar given all strategies would have the same reduction in total cost. The preferred strategy for each country is driven by opportunity costs and each jurisdiction’s priorities and willingness to pay.

Thresholds for cost-effectiveness worldwide vary; in the United States, the threshold is typically $100,000 while the threshold is generally considered to be around $50,000 CDN ($36,784 USD)^19^ in Canada and between 20,000-30,000 pounds ($25,245-$37,868 USD)^20^ in the United Kingdom. For lower income countries, the cost-effectiveness threshold is markedly lower ranging from $3-$8982 (2020 US dollars)^21^. In our analyses, we show that for countries of low to middle income, with willingness to pay thresholds between $350-$37,500, dexamethasone for severe COVID-19 infections would likely be the most favoured strategy based on current data. For very low-income countries, the price of dexamethasone may be unaffordable and so supportive care would be the preferred strategy.

COVID-19 has had disproportionate impacts on patients in the United States based on ethnic background, location and socioeconomic status. Patients of colour including African Americans, Latinx and Native Americans, immigrants, patients in rural settings and patients with lower socioeconomic status in the United States have had increased morbidity and mortality due to COVID-19^22–24^. Similar findings have been reported worldwide, especially in countries of lower socioeconomic status^25^. This situation is further complicated by the limited global supply of remdesivir and its considerable expense for some jurisdictions. There is a significant risk that patients who could most benefit from remdesivir (moderate severity) may not have access to remdesivir, which will likely further negatively impact people of colour, rural patients or patients with lower socioeconomic status. In our model, the potential benefit loss without remdesivir is 0.002 QALY or 0.66 days; however, this possible benefit is enormous in aggregate. Increased access to and lower cost of remdesivir will be critical to best treat these more marginalized patients.

There has been one published cost-effectiveness analysis of remdesivir looking at cost-effective threshold prices finding that a treatment course of remdesivir should be approximately $19,000 to be cost effective based on a willingness to pay of $100,000 and about $4700 based on a willingness to pay of $50,000, suggesting that a cost recovery price should be between $1000-1600^26^. This model takes a lifetime perspective and uses remdesivir for all patients. Our model has a few key differences. We have taken a 1-year time-horizon, used severity-stratified drug efficacy based on current clinical trials data to better reflect the differences between moderate and severe infections, and utilized Medicare rates which may underestimate costs in the United States. In our analysis, we directly compare remdesivir versus dexamethasone for all hospitalized patients, as well as remdesivir for moderate and dexamethasone for severe infections to reflect current pragmatic treatment approaches based on the two available trials; we do not combine dexamethasone and remdesivir as treatment as there is no data for this. When we compare only remdesivir for moderate and severe infections versus best supportive care, we find an incremental cost-effectiveness ratio of $32,792/QALY at the current list price of remdesivir similar to the other published model.

There are several limitations to our model. First, our model is based on the available literature with relatively limited treatment randomized controlled trial outcome data available. Given that COVID-19 is an emerging disease with rapidly evolving literature, the assumptions in the model are subject to change, especially as the remdesivir report does not contain full 28-day mortality data and the dexamethasone study is only a preliminary report. Data regarding utility in COVID-19 does not yet exist and was extrapolated from similar experience with H1N1 and severe influenza. Our hospital costs were based on the Medicare price; in other centers, rates may be higher with private insurance. Last, we assumed that beyond the initial 28 days, there would be no further impact to health utility and mortality. Given COVID-19 was only first described in December 2019, 1-year data is not yet available about outcomes. Further, although there are some data regarding effects post infection^27^, the impact of COVID-19 after the initial infection is still to be determined. To mitigate the uncertainty, we performed a probability sensitivity analysis where the estimates of costs, utilities and probabilities were varied simultaneously over their distributions and found that remdesivir for moderate and dexamethasone for severe COVID-19 infections remained favoured. This model does not combine use of dexamethasone and remdesivir in individual patients as there is no published data for this strategy.

In summary, use of remdesivir for moderate infections and dexamethasone for severe infections emerged as the most cost-effective management for moderate-severe COVID-19 infections, and dexamethasone for severe infections was favoured with lower willingness to pay thresholds, although further data may change this conclusion. Additional information about the effect of remdesivir is required to better assess the cost-effectiveness of its use.

## Data Availability

Data inputs used for the model are available in Table 1.

## Data Availability

Data inputs used for the model are available in Table 1.

## Author Contributions

SEC: Conceptualization, analysis of data, writing original draft, final approval of the submitted manuscript.

RV: Interpretation of data, critical revision for important intellectual content, final approval of the submitted manuscript.

CB Interpretation of data, critical revision for important intellectual content, final approval of the submitted manuscript.

FMC: Critical revision for important intellectual content, final approval of the submitted manuscript.

LS: Conceptualization, interpretation of data, critical revision for important intellectual content, final approval of the submitted manuscript

## Competing Interests Statement

Dr. Congly reports grants from Gilead Sciences, Boehringer Ingelheim, Genfit, Allergan, and Sequana Medical and personal fees from Intercept Pharmaceuticals and Eisai outside the submitted work.

Drs. Varughese, Brown, Clement and Saxinger report no potential conflicts of interest.

This study was unfunded.

